# Comparative Analysis of three machine learning models for Early Prediction of Skeletal Class-III Malocclusion from Profile Photos

**DOI:** 10.1101/2022.07.26.22277593

**Authors:** Banu Kiliç, Tuğba Önal-Süzek, Selahattin Aksoy

**Author notes:** These authors contributed equally (Banu Kiliç, Selahattin Aksoy). Cite Aksoy S,Kılıç B,Önal-Süzek T(2022). “Comparative Analysis of Deep Learning and Machine Learning Models for Early Prediction of Skeleton Class-III Malocclusion from Profile Photos”.

## Abstract

The pre-adolescent growth period is the best time for the skeletal Class-III malocclusion treatment. Diagnosis and treatment during this period continue to be a complex orthodontic problem. Class-III malocclusion is complicated to treat with braces frequently requiring surgical intervention after a pubertal growth spurt. In addition, delayed recognition of the problem will yield significant functional, aesthetic, and psychological concerns. This study presents the first fully automated machine learning method to accurately diagnose Class-III malocclusion applied across mobile images, to the best of our knowledge. For this purpose, we comparatively evaluated three machine learning approaches: a deep learning algorithm, a machine learning algorithm, and a rule-based algorithm. We collected a novel profile image data set for this analysis along with their formal diagnosis from 435 orthodontics patients. The most successful method among the three was the machine learning method, with an accuracy of %76.

## 1. Introduction

Skeleton Class-III malocclusion is characterized by the sagittal developmental retardation and/or rearward positioning of the maxilla or the extreme development and/or forward positioning of the mandible [1]. Class-III malocclusion causing a concave profile causes aesthetic anxiety in the child and the parents [2]. The correction of the harmony between the mandible and maxilla can be provided by orthodontic/orthopedic appliances to be performed in early terms on the patients affecting the active sutures unclosed. Although the ideal age of treatment with orthopedic devices is preferably eight at the latest, it has also been reported that they can be effective up to 12 years of age [3]. It has been shown that early Class-III orthopedic treatment reduces the need for orthognathic surgery [4]. The American Association of Orthodontists (AAO) recommends that all children be checked by an orthodontist no later than age seven. However, the most common age group during which patients seek orthodontic treatment is 12 years and older [5]. Moreover, many children do not have access to an orthodontic pre-examination at seven and before. Advances in consumer electronics and portable communications systems, particularly mobile phones, have led to faster and less expensive approaches to developing Point of Care Diagnosis[6]. Number of mobile subscribers globally according to GSMA Intelligence 2021 data reached approximately 5.22 billion [7]. Triggered by the Covid-19 pandemic period, the tele-health industry and more specifically the mobile health industry has become a 100 billion dollar market by increasing five times since 2016 as of the end of 2021 [8]. There are several dental mobile applications in the current healthcare mobile application market [9]. Current applications are mainly developed for patient education about general dentistry [10], braces, Invisalign (Align Technology, San Jose, Calif), and oral health [11]. The commercial software Phimentum[12] claims using deep-learning for automatic landmark point detection on cephalometric images. Unlike Phimentum, our program has the advantage of not requiring cephalometric image input and/or an orthodontist’s supervised selection of landmark points can compute the probability of Class-III diagnosis without any other apparatus other than mobile phones.

Our previous study [13], implemented an unsupervised diagnostic method based on the angles computed for Turkish adult patients [14]. Our previous study’s disadvantage was that it was only applicable to Turkish adult patients. Our present study aimed to compare the accuracies of three alternative unsupervised machine learning approaches for malocclusion diagnosis. Best performing model of three will be adaptable to be trained on any given image set of any race or age rather than a fixed threshold determined for Turkish patients. For this purpose, we implemented and presented the accuracies of three methods; deeplearning algorithm, a machine learning algorithm, and a rule-based algorithm for an in-house image data set we collected from orthodontics patients.

## 2. Collection of Data

The data set created within the project’s scope consists of profile photos of patients visiting Orthodontics at Bezmialem Vakıf University Faculty of Dentistry. The Institutional Review Board approval (IRB# 54022451-050.05.04) was obtained from Bezmialem Vakıf University to use profile image data of patients who applied to Bezmialem Vakıf University in our project. The orthodontist with 20 years of orthodontics clinical experience diagnosed the patient images and classified them into three classes, Class-I, II, and III using Dolphin Imaging software (Version 11.95). The profile image taken for each patient is saved with an anonymized name in JPG/PNG format. Our anonymized dataset consists of 435 profile images and with their formal diagnosis from the orthodontics clinic.

## 3. Materials and Methods

Three alternative methods, namely the Rule-based, Deep Learning, and Machine Learning methods were implemented (Fig-1).

**Figure 1.**
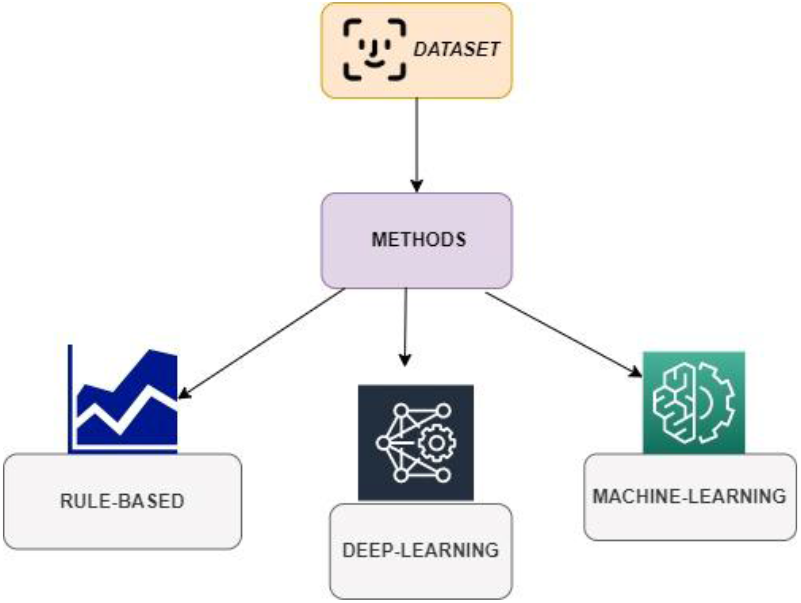
Flow-chart of the implemented pipeline

In this three methods we used mainly python programming language and it’s libraries scikit-learn for Machine Learning approach, OpenCV for image processing and pandas, seaborn etc. libraries for visualization and data analysis. In addition to that we used Tensorflow-Keras for deep learning approach. We used script base complier to run our python codes and other required python libraries.

### 3.1. Deep Learning Approach

For Deep Learning Method, firstly, the images were pre-processed, and then the deep learning model was created.

The images were first converted to gray-scale using the Python Open-CV library. Each image was scaled to 255×255 to standardize the images. Gaussian filtering was applied to scaled images using a 5×5 kernel matrix using OpenCV. Gaussian filtering (Gaussian Blur) was applied to reduce the noise and detail. Then, the median values were computed for each picture. With these median values, the contours of the profile images were extracted using OpenCV “Canny Edge detection” on each picture. Using dilation (Spreading and Expansion) and erosion operations pixels were added to the silhouette to make them more prominent. We applied dilation to expand the borders; thanks to this expansion, we enlarged the pixel groups and reduced the spaces between the pixels.

Next, the images were normalized using the min-max normalization formula (Fig-4), as all the images have a matrix with pixel values (0,255). As a result of this process, we converted our pictures into binary (0,1) form. Afterwards, we converted our matrix into a one-dimensional list by applying Flatten in order to use the 2D picture matrices we obtained in deep learning. As a result each picture’s silhouette, (Fig-3) was acquired in a single dimension.

**Figure 2.**
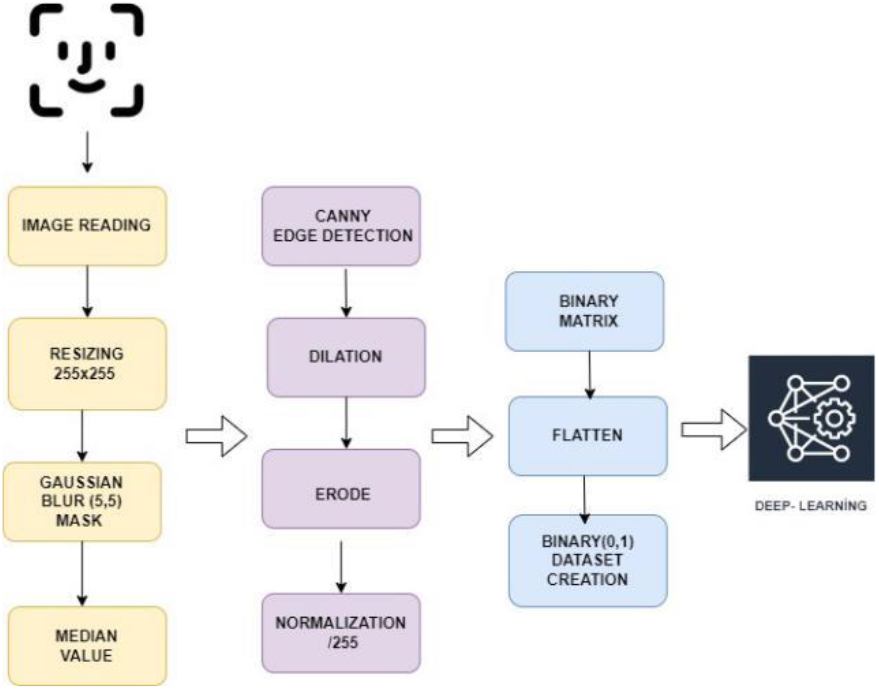
Image processing pipeline.

**Figure 3.**
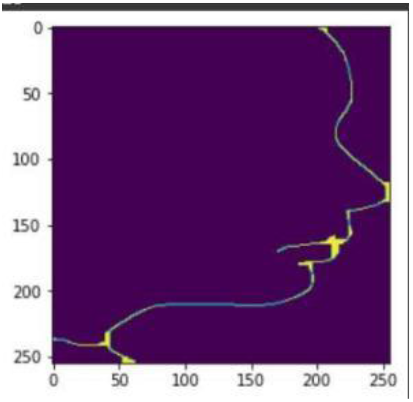
Silhouette output of the deep-learning pre-processing.

**Figure 4.**
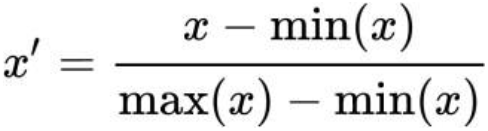
Normalization formula.

The images were split into 80% training and 20% test set. 80% of the images were used for training the deep-learning model. Our deep learning model used a 4-layer structure, an input layer, two hidden layers, and an output layer. Since we were aiming a binary classification, we used “Relu” as the activation function in our input layer, and we used the sigmoid function in the intermediate layers. We used “Adam” as the optimizer and “cross entropy” as the optimizer loss function.

After creating the training model, we tested our model with our independent test data. As a result, a deep-learning model with 70% accuracy and 64% sensitivity was created for the disease classification. As a higher accuracy was required for a clinical setting, we experimented with additional unsupervised methods that do not require a pre-set angle coefficient for diagnosis. To achieve this goal, we implemented the rule based method as the next step.

### 3.2 Rule Based Approach

The same re-sizing and pre-processing steps were carried out for the critical value method on the images. Using the Python face-alignment library [15], 68 facial landmark points were selected., Po’, Sn’, A’, Ls, Li, B, Pg’, and Gn’ were determined among these landmarks. After determining the other points requested by the orthodontist (Fig-6), the area between the Po’ point at ear level and the area surrounding the Li (Lower Lip) part green area in Fig-7) and the remaining area between Ls - Sn’-Po’ (red zone in Fig-7) Python OpenCV After masking, the ratio of the regions were computed in pixels.

**Figure 5.**
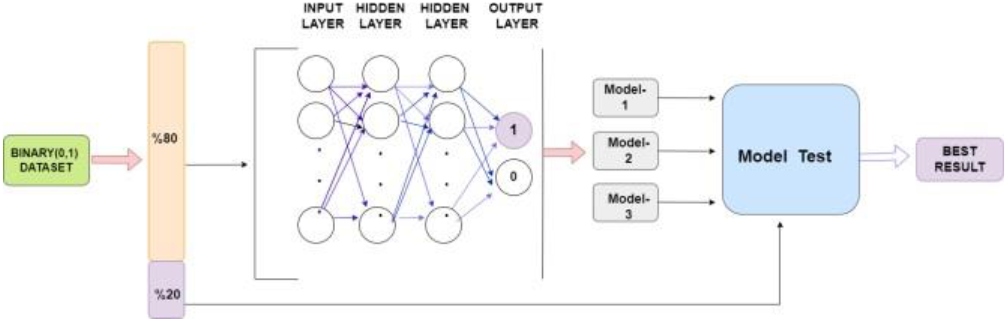
Deep learning model used.

**Figure 6.**
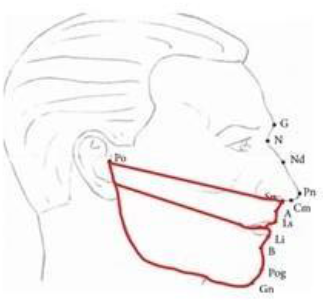
Two regions compared.

**Figure 7.** Two regions compared.

For the area ratios of all patients, a total of 435 images, 162 Class-I, 163 Class-II, and 110 Class-III, we first plotted the descriptive statistical analysis and visualization.

Histogram graphs were plotted to show the distribution of each class; Class-I, Class-II, and Class-III (Fig-8). As seen in the histograms there is no left or right skewness (Left-Right Skewed Distribution) and the data show a normal distribution

**Figure 8.**
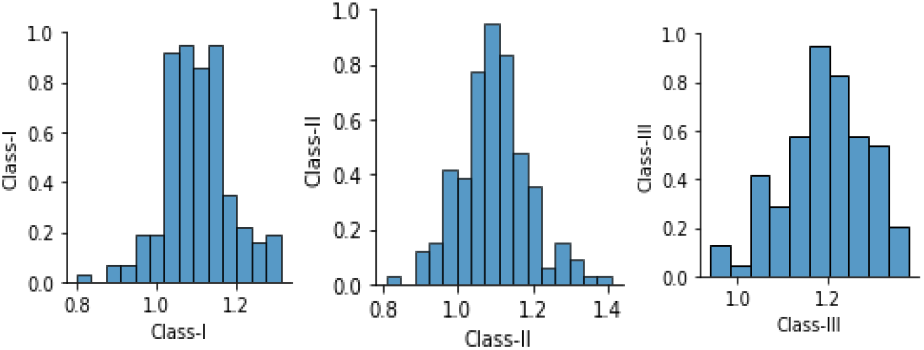
Histogram of each class showing the normalized number of samples.

The boxplots of the three classes in Fig-9 imply that the area ratios of Class-III patients were distinctly different from the other classes. Outliers in the boxplot were filtered and checked for misdiagnosis.

**Figure 9.**
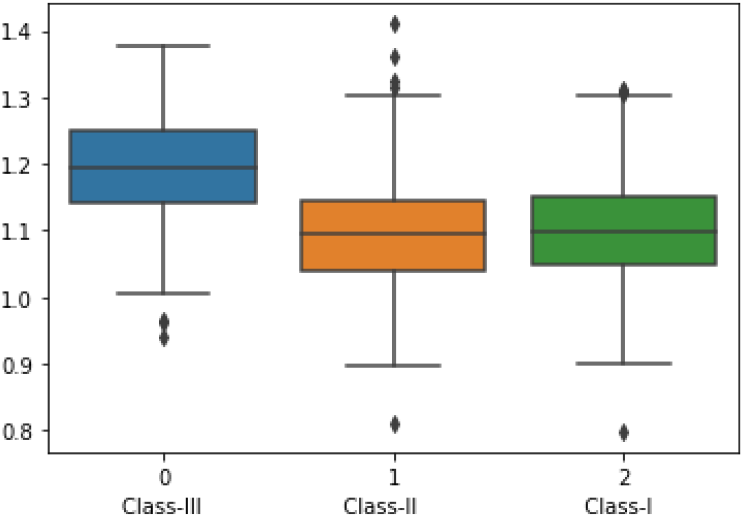
Boxplots of Class-I, Class-II, and Class-III.

In Fig-10, scatter plot of the patients also shows that Class-III patients have distinct area ratio values. As seen in the density plot in Fig-11, the area values showed a normal distribution for all classes yet the mean of the Class-III area ratio was higher than the other two classes.

**Figure 10.**
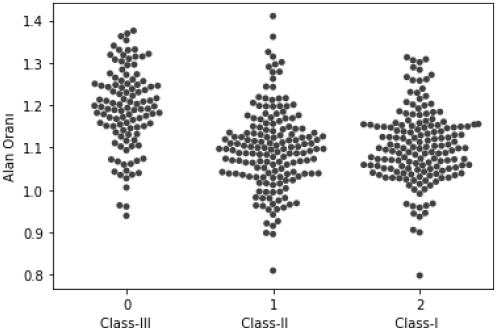
Scatter plot.

**Figure 11.**
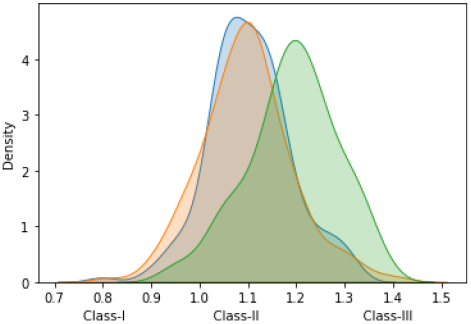
Density plot.

#### 3.2.1. Evaluation of Class-III vs not

Classification of area ratios for our goal, we designed binary classification experiment of Class-III and not Class-III. In this setting, 110 patients were Class-III and 325 patients were not Class-III (Fig-12).

**Figure 12.**
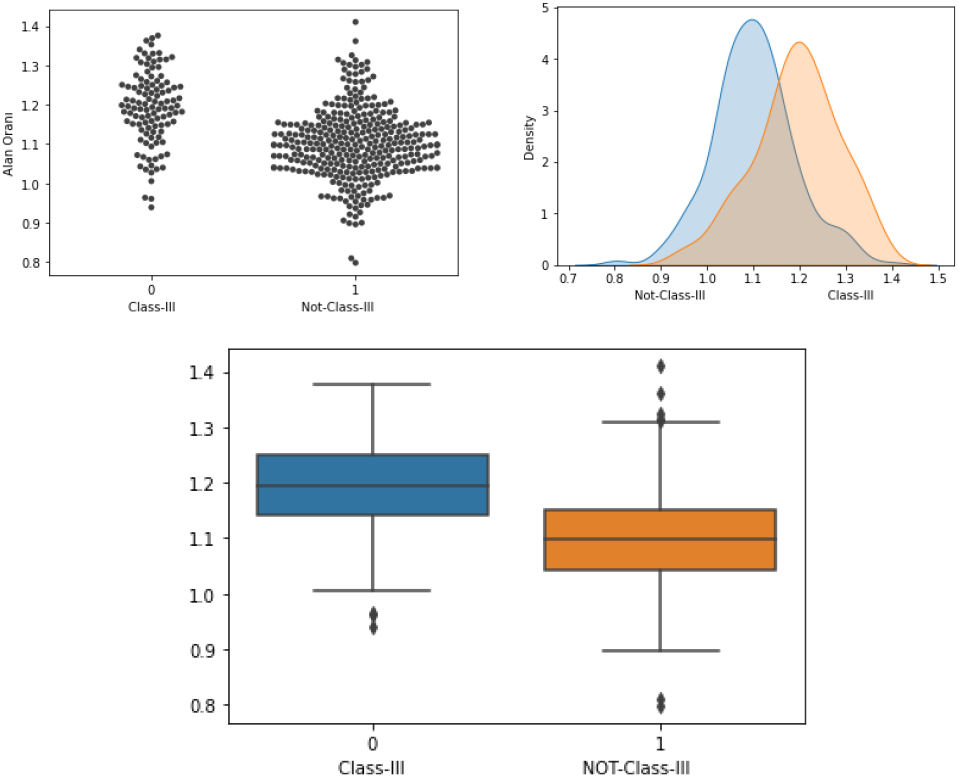
Distributions of Class-III and non-Class-III.

The area ratios of Class-III and non-Class-III followed a normal distribution (Figure 12-13).

**Figure 13.**
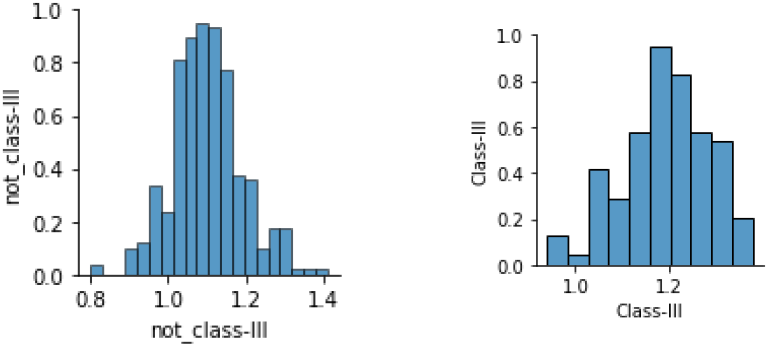
Histograms of Class-III and non-Class-III.

#### 3.2.2. Cleaning Up Outliers

Outliers were detected using the formula of Interquartile range (IQR) [Q1-1.5*IQR -Q3+1.5*IQR], we found 12 outliers and removed them from the dataset. We obtained a total of 423 patients. We repeated the analysis as in Fig-14.

**Figure 14.**
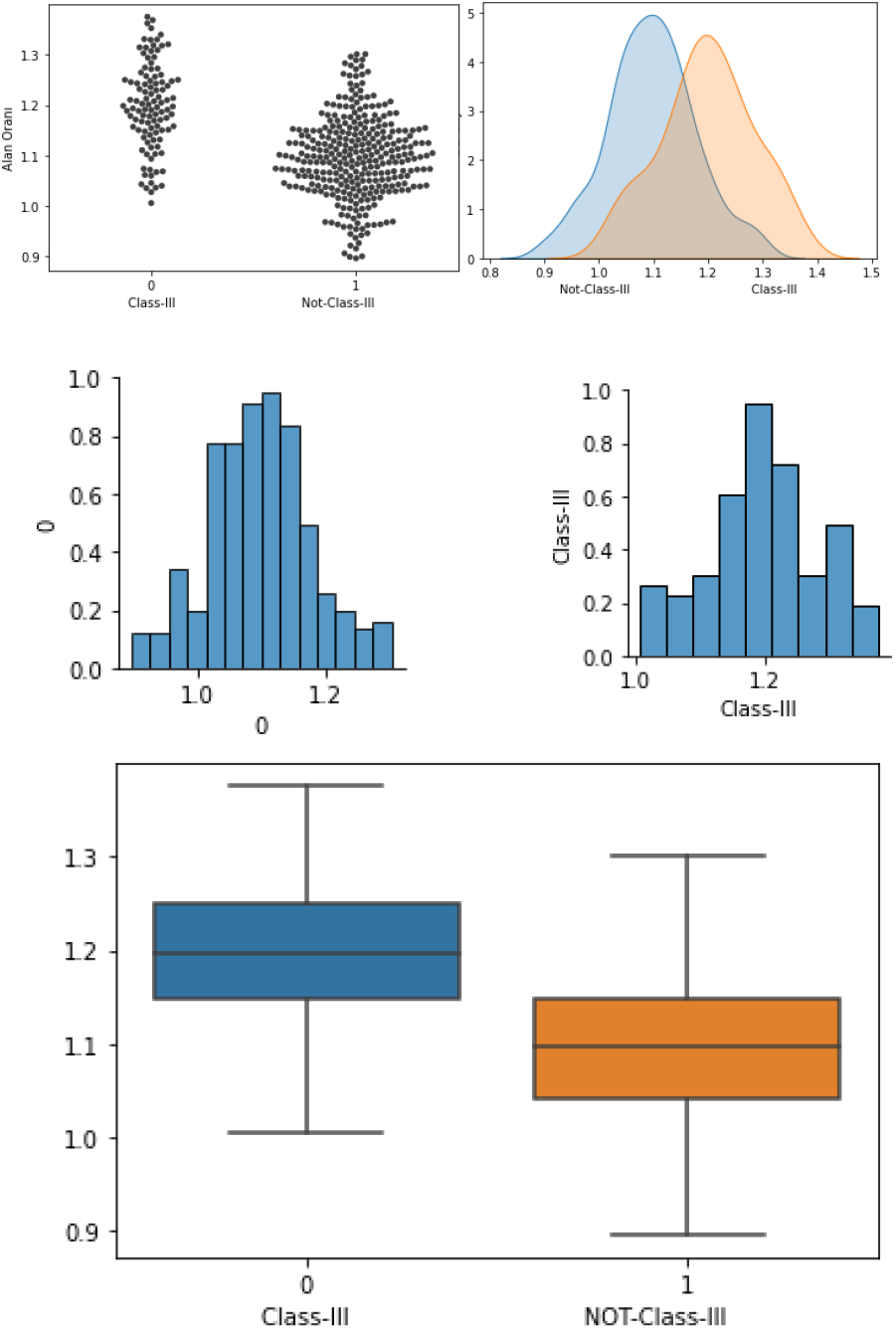
Plots of Class-III and non-Class-III after cleaning outliers.

#### 3.2.3 Experimental Setup

To check whether the distinct area ratio difference of Class-III is random or not, we have created an experimental setup (Fig-15) in the form of a critical value (threshold). In the experiment setup, we divided the Class-III and non-Class-III data into 5 parts with 5-fold separately, and the average of 4 parts of the data set was labeled training. The same process was repeated for the non-Class-III dataset. The resulting rule-based value was tested with the independent test dataset of Class-III and non-Class-III data. The process was repeated for the entire 5-fold. The classification was labeled not Class-III if the test data area ratio was greater than the critical value and Class-III if it was less than the critical value. As a result of classification

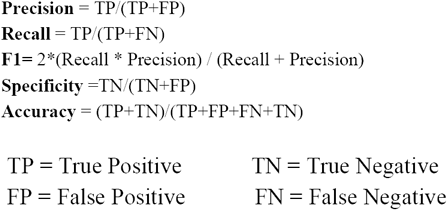

**Figure 15.**
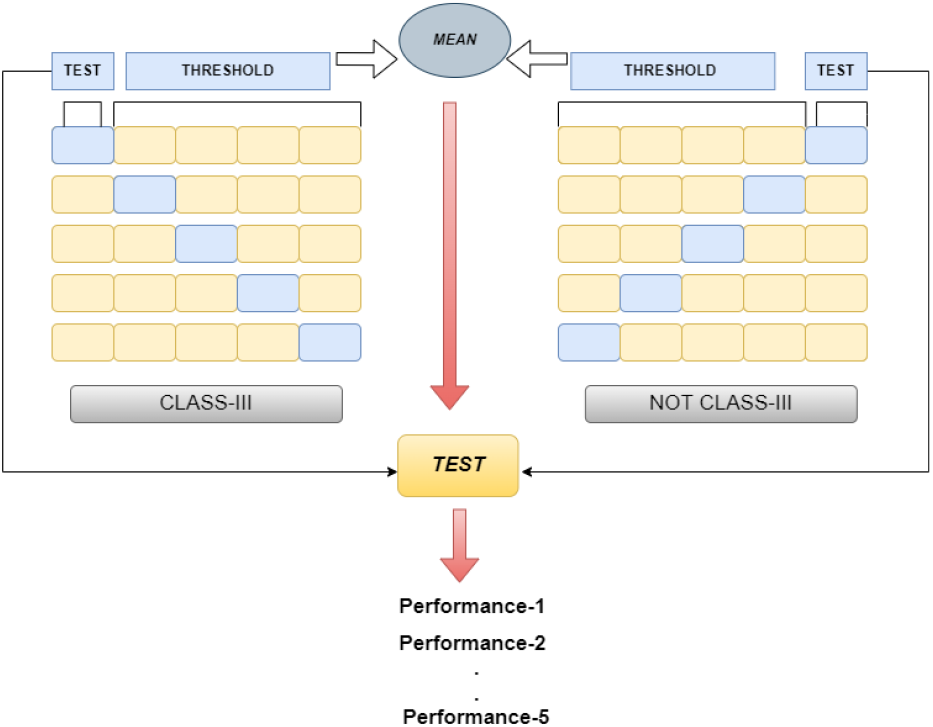
Experimental set up for classification.

#### 3.2.4. Results

We shuffled our experimental dataset 100 times to avoid bias, dividing the dataset 5-fold each time. The average and standard deviation of these 100 training shuffles were computed below.

#### 3.2.5. Results(without outliers)

The same calculation and method were also applied on the data without the outliers, which increased the performance by ∼%2.

After clearing the outliers we randomly shuffled our dataset each time, divided it into 5-Folds, repeated it 100 times, and calculated the average and standard deviation of these 100 training tests so that there would be no bias during training and testing.

According to the results of the area ratio rule based experiment, specific facial landmarks such as Po’, Sn’, A’, Ls, Li, B’, Pog’, Gn’, the area surrounding the Po’ point at the level of the Li (Lower Lip) and the remaining area between Li’ - Sn’ - Po’ (red zone in Fig-7) differs distinctly for skeletal Class-III malocclusion patients. The ratio method is an important distinguishing factor for Class-III classification; therefore, we decided to add it as a feature to our machine learning model.

### 3.3. Machine Learning Approach

The machine learning method was performed in two main steps.

3.3.1) Extraction of orthodontic features from images.

3.3.2) The data set was labeled and the machine learning method is applied.

#### 3.3.1. Feature Extraction from Images

For calculation of H-Angle, we used the Python face-alignment library [15]. Firstly soft tissue nasion(N’), soft tissue chin(Pg’) and upper lib(Ls) points (see Table 2) were computed. A line was drawn between Pg’ and N’ and a line between Pg’ and Ls, and the angle between the lines passing through these points was calculated as in Fig-17.

**Table 1.** Accuracy of the deep learning method.

|  |  |
| --- | --- |
| **Accuracy** | **0.7090909090909091** |
| **Precision** | **0.6470588235294118** |
| **F1** | **0.5789473684210527** |
| **Recall** | **0.5238095238095238** |
| **AUC** | **0.6736694677871147** |

**Table 2.** Descriptions of facial landmarks.

| Abbreviations | Definitions |
| --- | --- |
| N' | Soft tissue Nasion. Midpoint on the soft tissue contour of the base of the nasal root at the level of the frontonasal suture. |
| Ls | Labialis superior; the most anterior point of the upper lip. |
| Li | Labialis inferior; The most anterior point of the lower lip |
| Pg' | Soft tissue pogonion. The most anterior point on the mandible in the midline; the most anterior, prominent point on the chin. |
| Po' | Soft tissue porion : most superior point of the external acoustic meatus. |
| Pn | Pronasale; The tip of the nose. |
| Sn' | Soft tissue subnasal: The point at which thenasal septum merges with the uppercutaneous lip in the midsagittal plane. |
| A' | Soft tissue A point; The outer point of intersection between the A point horizontal line and the soft tissue. |
| B' | Soft tissue B point; The outer point of intersection between the B point horizontal line and the soft profile. |
| Soft Tissue Facial Plane | Line between soft tissue nasion to soft tissue pogonion |
| H line | Harmony line: drawn tangent to the soft tissue chin and the upper lip. |
| H angle | The angle formed between the softtissue facial plane line and the H line |

**Figure 16.**
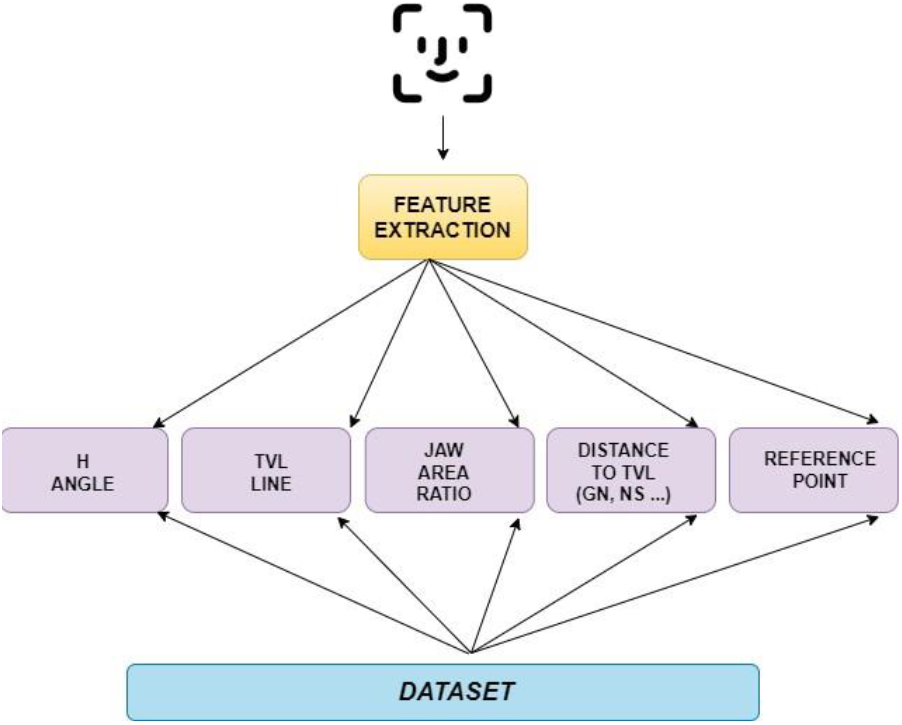
Extraction of orthodontic features from images.

**Figure 17.**
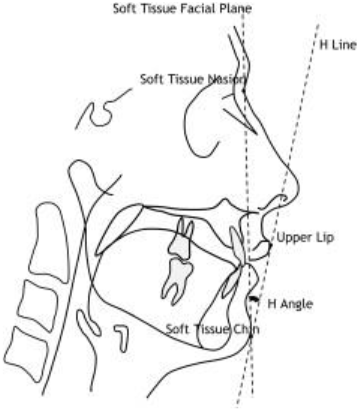
H-Angle of profile images.

To calculate the H angle in between, these points’ x and y coordinate values were extracted as a 2-dimensional plane. With these coordinates, the slopes of these two lines were calculated using the formula in Fig-18-b, and the angle H, which is the angle between the two lines, was calculated with the formula in Fig-18-a using these slopes.

**Figure 18.**
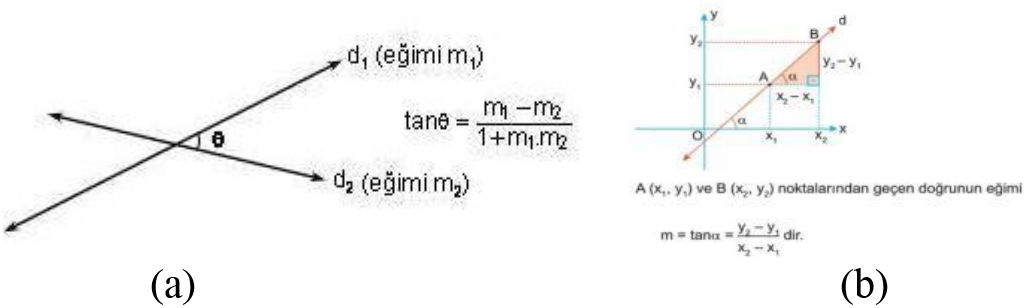
(a) Angle between 2 points, (b) slope of a line.

The threshold value computed in section 3.2(rule based method) was incorporated as an additional feature.

For TVL line feature, 68 reference points on the face were computed as x and y values by drawing a parallel line to the Sn (Subnasale) point and perpendicular to the transverse plane.

The distances of each point to the TVL line were computed, and distance ratios were calculated as taking the TVL line as a reference. We aimed for a correct diagnosis from profile pictures taken by any mobile phone, but general variables of the images such as proximity, distance, location, and angle were not standard.. To achieve higher accuracy with the nonstandard photos, we decided that our machine learning model should handle the lengths of the points from the TVL as they were scalable. For this purpose, we scaled all the profile photos by dividing the distances between the landmark points by a reference length. To pick the best reference length to scale by, we experimented with all paired combinations of 68 points. As a result, when the TVL was divided by the length between G and Gn, the most accurate distance to TVL scaling was obtained. The distances between the “Gn’”, “Sn’”, “A’”, “UL’”, “LL’”, “B’” and “Pog’” points on the face and the TVL were computed.

#### 3.3.2. Machine Learning Model

Firstly, we divided our data set into 75% training and 25% testing. Next, we selected the most appropriate model and parameter optimization on 75% of the training data. We calculated the correlations for the features, evaluated the features’ correlation against each other, and removed the “Pog’” data, as its correlation was greater than 0.80. No data was removed since we created our dataset by feature extraction from images.

Since the data set, we used in the machine learning method was not balanced, we created a balanced data set with random selection of the negative class and kept the entire positive class. We compared the results by repeating the experiment twice for our balanced (Table-6) and unbalanced data sets (Table-7). In this experiment, we used 5-fold cross-validation for model selection and repeated this experiment 100 times on 18 models (Fig-20). We selected the model with the best results after the cross-validation. After hyperparameter optimization, the best parameters were selected. By creating our models according to these parameters, we tested them on the test data and selected the model with the best results.

**Table 3.** The result of the experiment (3.2.3) for the whole data set.

|  | Accuracy | Recall | Precision | F1 | Specificity |
| --- | --- | --- | --- | --- | --- |
| **Fold- 1** | 0.712644 | 0.636364 | 0.451613 | 0.528302 | 0.738462 |
| **Fold- 2** | 0.735632 | 0.727273 | 0.484848 | 0.581818 | 0.738462 |
| **Fold- 3** | 0.666667 | 0.772727 | 0.414634 | 0.539683 | 0.630769 |
| **Fold- 4** | 0.758621 | 0.863636 | 0.513514 | 0.644068 | 0.723077 |
| **Fold- 5** | **0.793103** | 0.727273 | 0.571429 | 0.640000 | 0.815385 |
| **Mean** | 0.733333 | 0.745455 | 0.487208 | 0.586774 | 0.729231 |
| **STD** | 0.042637 | 0.073855 | 0.053525 | 0.048532 | 0.058865 |

**Table 4.** Summary of the mean accuracies of 100 repeats.

|  | Accuracy | Recall | Precision | F1 | Specificity |
| --- | --- | --- | --- | --- | --- |
| **Mean** | 0.734597 | 0.749545 | 0.486601 | 0.58755 | 0.72953 |
| **STD** | 0.0351 | 0.0921 | 0.04608 | 0.0502 | 0.0502 |

**Table 5.** The result of the experiment (3.2.3) for the cleaned dataset.

|  | Accuracy | Recall | Precision | F1 | Specificity |
| --- | --- | --- | --- | --- | --- |
| **Fold- 1** | 0.75581 | 0.81818 | 0.51428 | 0.63157 | 0.73437 |
| **Fold- 2** | 0.72941 | 0.72727 | 0.48484 | 0.58181 | 0.73015 |
| **Fold- 3** | 0.75000 | 0.57142 | 0.50000 | 0.53333 | 0.80952 |
| **Fold- 4** | 0.69047 | 0.71428 | 0.42857 | 0.53571 | 0.68254 |
| **Fold- 5** | 0.80952 | 0.80952 | 0.58620 | 0.68000 | 0.80952 |
| **Mean** | **0.74704** | 0.72813 | 0.50278 | 0.59248 | 0.75322 |
| **STD** | 0.03873 | 0.08886 | 0.050850 | 0.05660 | 0.04944 |

**Table 6.** Summary of the mean accuracies of 100 repeats.

|  | Accuracy | Recall | Precision | F1 | Specificity |
| --- | --- | --- | --- | --- | --- |
| **Mean** | 0.74869 | 0.75095 | 0.50363 | 0.59984 | 0.74793 |
| **STD** | 0.04788 | 0.09448 | 0.06632 | 0.06432 | 0.05978 |

**Table 7.** Balanced dataset results

|  | Accuracy | Precision | Recall | F1 | AUC |
| --- | --- | --- | --- | --- | --- |
| LR | 0.759<br>(0.071) | 0.753<br>(0.084) | 0.781<br>(0.115) | 0.761<br>(0.076) | 0.841<br>(0.065) |
| RC | 0.765<br>(0.069) | 0.754<br>(0.080) | 0.796<br>(0.101) | 0.770<br>(0.069) | 0.842<br>(0.065) |
| SGD | 0.681<br>(0.094) | 0.739<br>(0.150) | 0.684<br>(0.260) | 0.658<br>(0.144) | 0.836<br>(0.073) |
| PAC | 0.604<br>(0.097) | 0.689<br>(0.200) | 0.692<br>(0.335) | 0.595<br>(0.171) | 0.822<br>(0.073) |
| KNNC | 0.742<br>(0.068) | 0.729<br>(0.076) | 0.779<br>(0.103) | 0.748<br>(0.069) | 0.800<br>(0.069) |
| DT | 0.656<br>(0.075) | 0.661<br>(0.092) | 0.641<br>(0.120) | 0.644<br>(0.087) | 0.653<br>(0.076) |
| ETC | 0.653<br>(0.082) | 0.660<br>(0.096) | 0.652<br>(0.123) | 0.651<br>(0.088) | 0.652<br>(0.084) |
| L-SVC | 0.754<br>(0.070) | 0.754<br>(0.085) | 0.764<br>(0.126) | 0.752<br>(0.077) | 0.837<br>(0.065) |
| SVC | 0.765<br>(0.071) | 0.759<br>(0.085) | 0.785<br>(0.104) | 0.767<br>(0.073) | 0.836<br>(0.067) |
| NB | 0.732<br>(0.069) | 0.795<br>(0.108) | 0.645<br>(0.128) | 0.701<br>(0.086) | 0.802<br>(0.074) |
| ABC | 0.682<br>(0.075) | 0.685<br>(0.086) | 0.683<br>(0.117) | 0.678<br>(0.081) | 0.730<br>(0.081) |
| BC | 0.722<br>(0.071) | 0.743<br>(0.093) | 0.685<br>(0.117) | 0.703<br>(0.087) | 0.783<br>(0.076) |
| RF | 0.773<br>(0.070) | 0.788<br>(0.090) | 0.760<br>(0.115) | 0.765<br>(0.079) | 0.829<br>(0.069) |
| ETSC | 0.773<br>(0.069) | 0.788<br>(0.085) | 0.751<br>(0.105) | 0.765<br>(0.074) | 0.824<br>(0.069) |
| GPC | 0.769<br>(0.070) | 0.736<br>(0.075) | 0.847<br>(0.091) | 0.784<br>(0.066) | 0.815<br>(0.072) |
| GBC | 0.728<br>(0.070) | 0.735<br>(0.085) | 0.726<br>(0.111) | 0.723<br>(0.075) | 0.790<br>(0.072) |
| LDA | 0.730<br>(0.072) | 0.736<br>(0.084) | 0.726<br>(0.112) | 0.725<br>(0.078) | 0.815<br>(0.070) |
| QDA | 0.657<br>(0.068) | 0.757<br>(0.125) | 0.482<br>(0.151) | 0.571<br>(0.115) | 0.756<br>(0.076) |

**Table 8.** Unbalanced dataset results.

|  | Accuracy | Precision | Recall | F1 | AUC |
| --- | --- | --- | --- | --- | --- |
| LR | 0.825<br>(0.037) | 0.722<br>(0.114) | 0.510<br>(0.106) | 0.591<br>(0.095) | 0.859<br>(0.047) |
| RC | 0.816<br>(0.032) | 0.747<br>(0.125) | 0.423<br>(0.106) | 0.531<br>(0.099) | 0.859<br>(0.047) |
| SGD | 0.776<br>(0.100) | 0.638<br>(0.175) | 0.581<br>(0.224) | 0.570<br>(0.106) | 0.852<br>(0.050) |
| PA | 0.661<br>(0.220) | 0.579<br>(0.299) | 0.455<br>(0.362) | 0.372<br>(0.189) | 0.845<br>(0.056) |
| KN | 0.794<br>(0.042) | 0.616<br>(0.108) | 0.519<br>(0.117) | 0.556<br>(0.095) | 0.772<br>(0.060) |
| DT | 0.733<br>(0.049) | 0.473<br>(0.102) | 0.491<br>(0.123) | 0.475<br>(0.098) | 0.654<br>(0.065) |
| ETC | 0.725<br>(0.051) | 0.458<br>(0.104) | 0.469<br>(0.126) | 0.451<br>(0.104) | 0.638<br>(0.063) |
| L-SVC | 0.824<br>(0.035) | 0.733<br>(0.117) | 0.483<br>(0.108) | 0.577<br>(0.093) | 0.858<br>(0.047) |
| SVC | 0.819<br>(0.033) | 0.729<br>(0.121) | 0.466<br>(0.108) | 0.560<br>(0.096) | 0.853<br>(0.046) |
| NB | 0.794<br>(0.047) | 0.629<br>(0.126) | 0.517<br>(0.134) | 0.553<br>(0.097) | 0.810<br>(0.059) |
| ABC | 0.772<br>(0.041) | 0.560<br>(0.103) | 0.480<br>(0.119) | 0.510<br>(0.097) | 0.768<br>(0.068) |
| BC | 0.779<br>(0.042) | 0.597<br>(0.116) | 0.441<br>(0.113) | 0.499<br>(0.098) | 0.784<br>(0.057) |
| RF | 0.794<br>(0.037) | 0.624<br>(0.102) | 0.471<br>(0.114) | 0.525<br>(0.097) | 0.814<br>(0.054) |
| ETS C | 0.792<br>(0.038) | 0.617<br>(0.107) | 0.467<br>(0.117) | 0.528<br>(0.095) | 0.802<br>(0.054) |
| GPC | 0.810<br>(0.039) | 0.680<br>(0.122) | 0.485<br>(0.108) | 0.558<br>(0.097) | 0.819<br>(0.054) |
| GB | 0.788<br>(0.041) | 0.606<br>(0.110) | 0.482<br>(0.116) | 0.530<br>(0.098) | 0.805<br>(0.055) |
| LD | 0.814<br>(0.040) | 0.689<br>(0.121) | 0.499<br>(0.109) | 0.571<br>(0.098) | 0.842<br>(0.050) |
| QD A | 0.759<br>(0.036) | 0.560<br>(0.201) | 0.253<br>(0.151) | 0.326<br>(0.145) | 0.783<br>(0.062) |

**Table 9.**
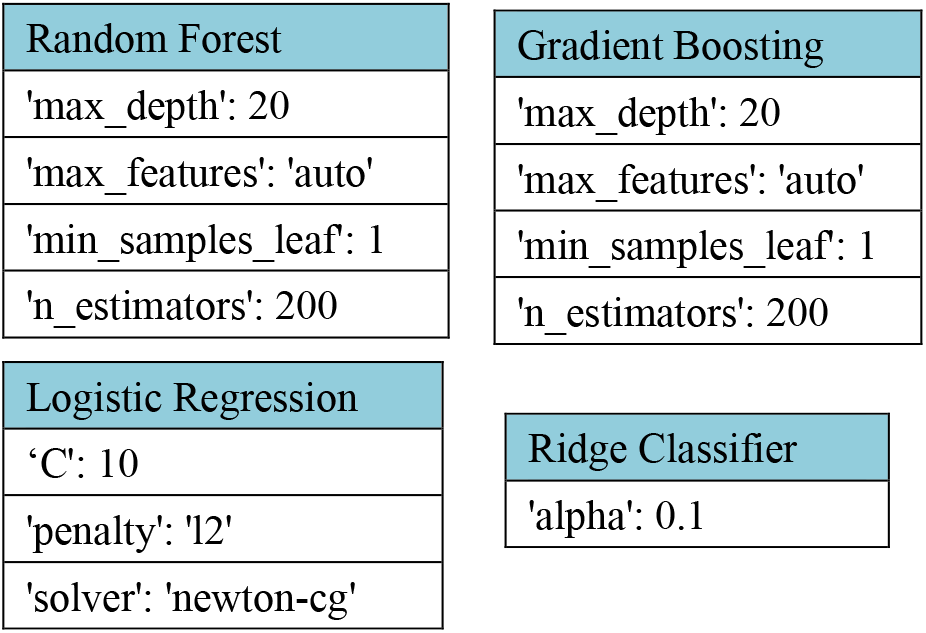
Hyperparameter optimization results for four models.

**Table 10:** Balanced data test results for each model.

| Random Forest Classifier |  |
| --- | --- |
| Accuracy | 0.6727272727272727 |
| Precision | 0.6666666666666666 |
| F1 | 0.689655172413793 |
| Recall | 0.7142857142857143 |
| AUC | 0.6719576719576721 |
| Gradient Boosting Classifier |  |
| Accuracy | 0.7636363636363637 |
| Precision | 0.7586206896551724 |
| F1 | 0.7719298245614034 |
| Recall | 0.7857142857142857 |
| AUC | 0.7632275132275131 |
| Ridge Classifier |  |
| Accuracy | 0.7636363636363637 |
| Precision | 0.7419354838709677 |

| **F1** | **0.7796610169491526** |
| --- | --- |
| **Recall** | **0.8214285714285714** |
| **AUC** | **0.7625661375661377** |
| Linear SVC |  |
| Accuracy | 0.7636363636363637 |
| Precision | 0.7586206896551724 |
| F1 | 0.7719298245614034 |
| Recall | 0.7857142857142857 |
| AUC | 0.7632275132275131 |
| Logistic Regression |  |
| Accuracy | 0.7636363636363637 |
| Precision | 0.7419354838709677 |
| F1 | 0.7796610169491526 |
| Recall | 0.8214285714285714 |
| AUC | 0.7625661375661377 |

**Table 11:** Unbalanced data test results for each model

| Logistic Regression |  |
| --- | --- |
| Accuracy | 0.7981651376146789 |
| Precision | 0.625 |
| F1 | 0.576923076923077 |
| Recall | 0.5357142857142857 |
| AUC | 0.7123015873015872 |
| Random Forest Classifier |  |
| Accuracy | 0.7981651376146789 |
| Precision | 0.6363636363636364 |
| F1 | 0.56 |
| Recall | 0.5 |
| AUC | 0.7006172839506173 |
| Gradient Boosting Classifier |  |
| Accuracy | 0.8256880733944955 |
| Precision | 0.68 |
| F1 | 0.6415094339622641 |
| Recall | 0.6071428571428571 |
| AUC | 0.7541887125220458 |
| Ridge Classifier |  |
| Accuracy | 0.8348623853211009 |
| Precision | 0.7777777777777778 |
| F1 | 0.6086956521739131 |
| Recall | 0.5 |
| AUC | 0.7253086419753088 |
| Linear SVC |  |
| Accuracy | 0.8165137614678899 |
| Precision | 0.6818181818181818 |
| F1 | 0.6 |
| Recall | 0.5357142857142857 |
| AUC | 0.724647266313933 |
| Extra Trees Classifier |  |
| Accuracy | 0.8348623853211009 |
| Precision | 0.75 |
| F1 | 0.6250000000000001 |
| Recall | 0.5357142857142857 |
| AUC | 0.7369929453262786 |

**Figure 19.**
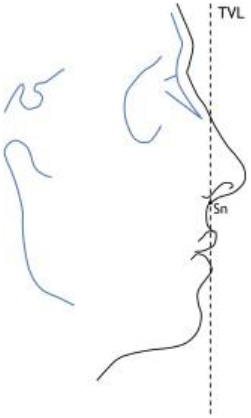
True Vertical Line (TVL).

**Figure 20.**
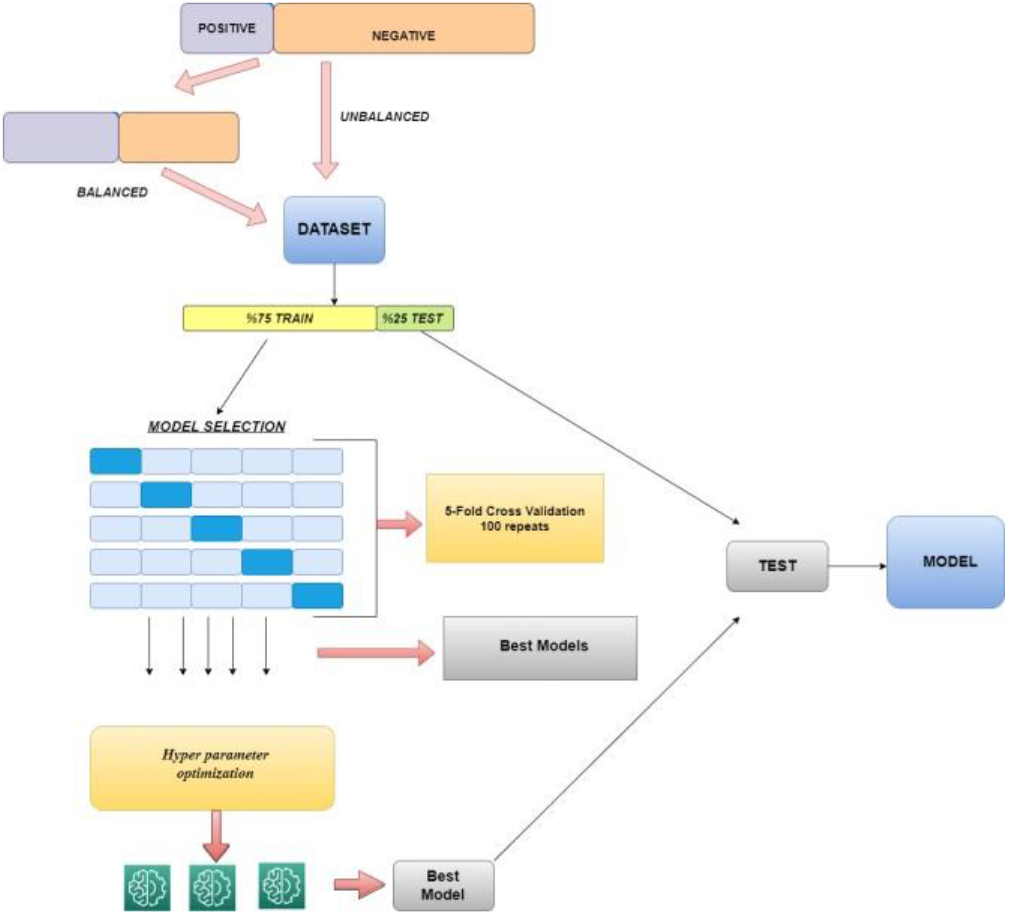
Experimental set-up for Machine Learning.

As the Random Forest, Logistic regression, Ridge Classifier, Extra Trees Classifier, Linear SVM performed best, and we optimized the parameters on these five models to select the most optimal parameters. In addition, we applied parameter optimization on a few more models with close cross validation performances.

#### 3.3.3. Hyper parameter optimization

We made hyperparameter optimization over the selecting models.

#### 3.3.4. Testing of Critical Models

Although the unbalanced dataset resulted in higher accuracy, F1 and precision were low. Therefore, balanced dataset’s accuracy, F1 and recall score more represent the actual production accuracy. The best result for the balanced machine learning model was Logistic Regression with 76% accuracy, 74% precision and 77% F1, 82% recall, and AUC of 76%.

## 4. Conclusion

This study presents the first fully automated machine learning method to accurately diagnose Class-III malocclusion applied across mobile images, to the best of our knowledge. For this purpose, we comparatively evaluated three machine learning approaches. We achieved 70% accuracy with the deep learning method, 74% accuracy with the area ratio method and 76% success with the machine learning method.

All three of our methods are flexible to be adapted to any dynamic training set of profile images of any ethnicity and age. Our next goal is to integrate our most accurate learning model among these three into a mobile application for parents and pediatricians seeking a second opinion on whether to reach out to an orthodontist at an early stage of developmental bone growth with a warning of Class-III malocclusion risk.

## Data Availability

All data produced in the present study are available upon reasonable request to the authors.

## 5. Acknowledgment

We want to thank Gül Sude Demircan, who developed the previous prototype, and Tülay Sevinç, who assisted in collecting the patient images and the consent forms.

## Appendix

|  |  |
| --- | --- |
| LR | Logistic Regression |
| RC | Ridge Classifier |
| SGD | SGD Classifier |
| PA | Passive Aggressive Classifier |
| KN | K Neighbors Classifier |
| DT | Decision Tree Classifier |
| ETC | Extra Tree Classifier |
| L-SVC | Linear SVC |
| SVC | SVC |
| NB | Gaussian NB |
| ABC | Ada Boost Classifier |
| BC | Bagging Classifier |
| RF | Random Forest Classifier |
| ETSC | Extra Trees Classifier |
| GPC | Gaussian Process Classifier |
| GB | Gradient Boosting Classifier |
| LD | Linear Discriminant Analysis |
| QDA | Quadratic Discriminant Analysis |

